# Interrogating structural inequalities in COVID-19 Mortality in England and Wales

**DOI:** 10.1101/2021.02.15.21251771

**Authors:** Gareth J Griffith, George Davey Smith, David Manley, Laura D Howe, Gwilym Owen

## Abstract

**Background:** Numerous observational studies have highlighted structural inequalities in COVID-19 mortality in the UK. Such studies often fail to consider the complex spatial nature of such inequalities in their analysis, leading to the potential for bias and an inability to reach conclusions about the most appropriate structural levels for policy intervention.

**Methods:** We use publicly available population data on COVID-19 related- and all-cause mortality between March and July 2020 in England and Wales to investigate the spatial scale of such inequalities. We propose a multiscale approach to simultaneously consider four spatial scales at which processes driving inequality may act and apportion inequality between these.

**Results:** Adjusting for population age structure, number of care homes and residing in the North we find highest regional inequality in March and June/July. We find finer-grained within-region increased steadily from March until July. The importance of spatial context increases over the study period. No analogous pattern is visible for non-COVID mortality. Higher relative deprivation is associated with increased COVID-19 mortality at all stages of the pandemic but does not explain structural inequalities.

**Conclusions:** Results support initial stochastic viral introduction in the South, with initially high inequality decreasing before the establishment of regional trends by June and July, prior to reported regionality of the “second-wave”. We outline how this framework can help identify structural factors driving such processes, and offer suggestions for a long-term, locally-targeted model of pandemic relief in tandem with regional support to buffer the social context of the area.

**Key Messages:**

- Regional inequality in COVID-19 mortality declined from an initial peak in April, before increasing again in June/July.
- Within-region inequality increased steadily from March until July.
- Strong regional trends are evident in COVID-19 mortality in June/July, prior to wider reporting of regional differences in “second wave”.
- Analogous spatial inequalities are not present in non-COVID related mortality over the study period.
- These inequalities are not explained by age structure, care homes, or deprivation.

## Background

Inequality in COVID-19 outcomes has been a matter of keen interest in the UK since the early stages of the pandemic (Bambra et al. 2020; Public Health England 2020). The “second wave” of Sars-COV-2 infection and subsequent COVID-19 related illness across the UK has been characterised by a strong regionality and an emergent North-South divide in mortality which both reflected and exacerbated existing social inequalities (Bambra, Norman, and Johnson 2020; Dorling and Davey Smith 2020).

An enormous literature has evolved over the course of the pandemic on individual predictors of COVID-19 related outcomes (e.g. Chadeau-Hyam et al., 2020; Williamson et al., 2020). A smaller body of work has also emerged looking the area-level predictors of COVID-19 outcomes and exposures (e.g. Harris 2020; Jay et al. 2020; Sundaram et al. 2020). Such spatial investigations have, however, largely focused on COVID-19 case status which is subject to temporally varying testing priorities (Griffith et al. 2020). As testing is more likely for severely ill or hospitalised individuals, we assume data on COVID-19 related mortality is less likely to be severely biased than case numbers as, conditional on life-threatening conditions, COVID-19 status will be more accurately reported amongst those in critical care.

We conceptualise COVID-19 mortality as a function of two processes: the risk of infection, and the risk of death given infection. Each of these is unlikely to have a uniform spatial distribution. Spatial analyses of aggregated mortality data are critical to epidemiological triangulation efforts as they give (fallible) measures of population parameters not subject to the same selection processes as individual level analyses (Lawlor, Tilling, and Smith 2016). We suggest predictors of aggregate mortality may primarily reflect risk of infection, as evidence does not suggest viral evolution has affected disease prognosis (du Plessis et al. 2021; Volz et al. 2021) and restricted migration patterns (given suppression measures) are unlikely to explain short term monthly differences within an age-adjusted population structure.

Investigation into COVID-19 inequalities has largely been restricted to analyses at a single spatial scale, predominantly Governmental Office Region (n=10 in England and Wales) (e.g. Northern Health Science Alliance, 2020) or smaller Middle-Layer Super Output Areas (MSOAs; n=7 201 in England and Wales) (e.g. Daras et al., 2020; Davies et al., 2020; Harris, 2020). However, limited spatial scope may bias results of such analyses as, when higher level clustering is omitted from a model, higher level phenomena are necessarily expressed at the included spatial scale (Tranmer and Steel 2001). Restricting policy relevant analysis to a single spatial scale risks overstating the importance of the included scale and missing structural exposures at which to target interventions and resources^1^. We provide a simple vignette outlining this problem in Box 1. Thus, the spatial nature of processes driving inequalities in COVID-19 mortality are currently under-investigated. To better understand area-level mortality differences, we must interrogate regional COVID-mortality inequality net of lower-level variation, and investigate at what stage of the pandemic it emerged.

### Box 1

The degree of spatial inequality which exists with respect to a given phenomenon depends on the spatial scale at which the question is asked (Jones et al, 2018, Manley et al, 2018). This is often referred to as the Modifiable Area Unit Problem (MAUP). A guided explanation of this is provided in Figure 1 below.

**Figure 1:**
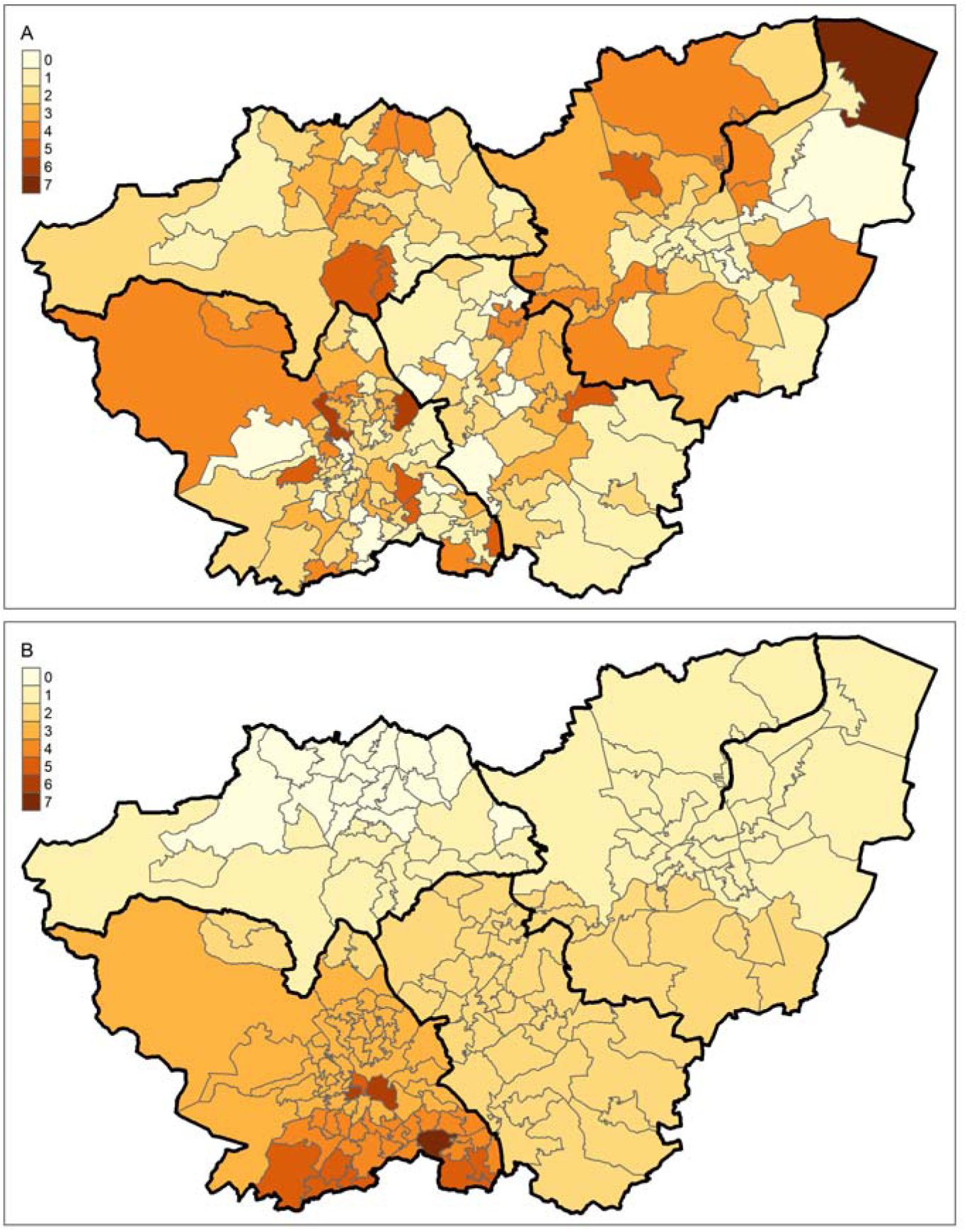
A map of South Yorkshire MSOAs, with 4 larger LADs highlighted by darkened bounds. Fig 1a shows a hypothetical scenario in which the large majority of inequality is truly between-MSOA, within-LAD. Fig 1b shows a hypothetical scenario in which the large majority of inequality is truly between-LAD, with very little within-LAD variation.

The map displays South Yorkshire. The smaller areal units are Middle-Layer Super Output Areas (MSOAs), and the areas outlined in thicker black lines are Local Authority Districts (LADs). In a single-level model of MSOA-mortality (e.g. Davies et al., 2020), the two maps below are identical. More explicitly, Poisson overdispersion (and residual error variation in linear regression) is assumed to be spatially random. This assumption is commonly violated, as it is in Figure 1B, where residuals are clearly spatially structured.

To meaningfully decompose this variation, we must consider both between- and within-interpretations of inequality. In Figure 1A, the majority of variation is within-LAD, between-MSOA. Net of any higher-level geography - this is close to spatially random, and we may assume that any spatial processes occurring in the data are at the MSOA-level. In Figure 1B, however, most variation is between-LAD, with very low between-MSOA, within-LAD variation.

If we only take account of the MSOA clustering (for instance by only including MSOA mortality counts) we would conclude, consistently across both plots, that MSOA is the level at which spatial processes are most likely to be occurring. Thus, in the scenario in Figure 1B we would fail to recognise that LAD-context may inform a structural exposure. However, if we simultaneously account for both MSOA and LAD, we can investigate the relative importance of each. Interrogating the relative importance of multiple geographical levels may better inform the likely exposures driving spatial inequalities, and hence inform potential interventions.

### Overview

In this article we use publicly available MSOA-level mortality for the period of March to June 2020 to unpack the geographical scale at which inequality in COVID-19 mortality was most keenly experienced. We rerun all analyses using non-COVID related mortality figures to contextualise inference about spatial processes unique to COVID-19. Having established area-level differences, we explore area level characteristics which may explain these inequalities.

We structure our analysis via the 3 following research questions:

1. What is the spatial scale at which mortality inequality (both COVID-19 related, and non-COVID-19 related) is most strongly expressed between March and July 2020?
2. Is spatial patterning consistent over the study period?
3. To what extent are these inequalities explained by local-area deprivation?

## Methods

### Data

All data used for this analysis are publicly available from the Office for National Statistics (ONS), Environmental Systems Research Institute (ESRI) or derived datasets made available in existing, published research.

### Outcome

Mortality data are counts of MSOA-level mortality from COVID-19 and non-COVID sources (Office for National Statistics 2020). These data provide provisional counts of numbers of deaths involving COVID-19 (defined as COVID-19 being mentioned on the death certificate) for each MSOA in England and Wales between the dates of 1^st^ March and 31^st^ July 2020. They also provide a total count of deaths from which the number of non-COVID-19 deaths can be calculated.

### Structural Levels

Investigating higher level spatial inequalities requires specification of plausible spatial scales at which mortality inequalities may be expressed. We select four analytical levels, chosen to represent plausible scales for spatial process, and also pragmatic levels for intervention.

The lowest level unit is the MSOA, the level at which the mortality counts are reported. We take the three administrative structures: MSOAs, Local Authority Districts (LADs) and Governmental Office Regions (GORs) as an intuitive representation of multiscale processes consistent with administrative boundaries. MSOAs (n=7201) are perfectly nested within LADs (n=348). Each LAD is also nested perfectly within one GOR (n=10).

As LADs and GORs differ greatly in size, we wanted to include a level which could capture processes between these two scales. As such, each MSOA is mapped onto a single Sustainable Transformation Partnership (STP, n=49). STP is chosen as spatially it sits between regional and LAD-level, and is of direct policy relevance for NHS governance and local councils (NHS 2016). As STP boundaries are defined by NHS commissioning and not administrative boundaries, MSOAs do not map perfectly onto them. There are 17 MSOAs which span STP borders, for each of these instances, we assign each MSOA to the STP within which the majority of its constituent Lower-Layer Super Output Areas lie. Finally, due to the spatial heterogeneity of LADs we also rerun models using ONS Travel to Work Areas (TTWAs, see Supplementary Analysis 1). TTWAs are more spatially consistent, and capture areas approximate labour markets, as areas in which the majority of population both live and work (Office for National Statistics 2016).

### Covariates

Older age has been established as the strongest individual predictor of COVID-19-specific mortality (Bhaskaran et al. 2021; Spiegelhalter 2020; Williamson et al. 2020), with 74% of COVID-19 deaths in the period March-July being amongst those over 75 (Office for National Statistics 2020). Care-home population has also been suggested the strongest area-level predictor of COVID-19 mortality (Davies et al. 2020; Harris 2020). We wish to estimate spatial inequalities net of these known risk factors, so adjust for them in all analyses. ONS mid-year population estimates for 2019 are used to account for MSOA age structure (ONS 2020). MSOA-level care home data is taken from the open-access ESRI COVID-19 dashboard Geolytix dataset which was made available for COVID researchers. Care homes are included as raw counts at MSOA level. North-South identifiers are generated by mapping GOR codes to the regional classification in Dorling, 2010.

We also wish to estimate inequality net of deprivation and so adjust for the Index of Mutliple Deprivation (IMD) as an indicator of the intersecting disadvantage shown to be associated with worse COVID-19 outcomes (Jay et al. 2020; Williamson et al. 2020). The IMD is country specific we would not be able to compare England and Wales, so we use the equivalised UK IMD (Abel, Barclay, and Payne 2016). This functions as an area-level indicator of disadvantage comparable across UK countries. Local healthcare was severely strained over the study period, so we decompose deprivation into between and within geographical components (i.e. regional average IMD, LAD minus region IMD, MSOA minus LAD IMD). This offers us insight into whether between-area differences in IMD are similarly predictive of excess mortality as within-area differences. Between- and within-elements of IMD are standardised. As these data were generated from 2015 IMD results for UK countries we rerun analyses excluding Wales and using 2019 IMD scores (Supplementary Analysis 2).

### Statistical analysis

To analyse multiscale inequality, we specify multilevel Poisson models with a geographically invariant, month-specific log-offset. The offset term indicates expected COVID-19 mortality and is calculated by taking the total COVID-19 mortality across England and Wales in each month and dividing by the total population. This is then multiplied by MSOA population to give an expected count were mortality rates spatially invariant. The same was done for non-COVID-19 related mortality. Models were stratified by mortality classification and were run for both COVID-19 and non-COVID-19 related mortality. Covariates were interacted with a categorical month term to allow effects to vary over time (full detail on model specification in Supplement).

Any overdispersion in random coefficients over and above this spatially consistent offset can be used to infer clustering, or spatial inequality. We summarise the variation in random coefficients using Median Rate Ratios (MRRs) (Austin et al. 2018; Jones et al. 2015). Transforming the variance from a typical interpretation (exp(σ)) allows for comparisons between standardised rates, without needing to consider effects in terms of a standard deviation increase in random effects.:

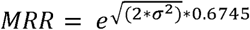

The MRR is interpreted as the median relative change in the mortality rate between randomly sampled pairs of lower-level units within a higher-level unit (Austin et al. 2018; Jones et al. 2015). For instance, an MSOA MRR of 1 would imply that there is no difference between risk of death in different MSOAs within the same LAD. An MSOA MRR of 2 implies that in general, comparing between MSOAs where all else is equal (i.e. within the same higher level unit, and balanced covariates), we would expect one of the MSOAs to have a mortality rate double that of the other. Where multiple higher-levels are included in a model, it is important to note that MRRs are estimated net of one another, such that inequality at each level is estimated separately to that in another.

Due to the non-hierarchical nesting of structural levels, MCMC estimation is used for all models. Restricted Iterative Generalised Least Squares estimates were taken as starting values. After a discarded burn-in of 50,000 iterations, all models were run for 500,000 iterations. Credible intervals for all estimated quantities are the 2.5^th^ and 97.5^th^ percentiles of posterior parameter distributions. All models were run in MLwiN v3.09.

## Results

Results are presented for COVID-19 and non-COVID-19 mortality. All models include fixed effect adjustment for being in the North, MSOA age structure, and number of care homes. Our first and second research questions regard the spatial and temporal variation in relative mortality risk over the study period and the relative contribution of each spatial scale. Monthly mortality counts are provided in Supplementary Table 1. Results for the first pair of models are presented in Figure 2.

**Figure 2:**
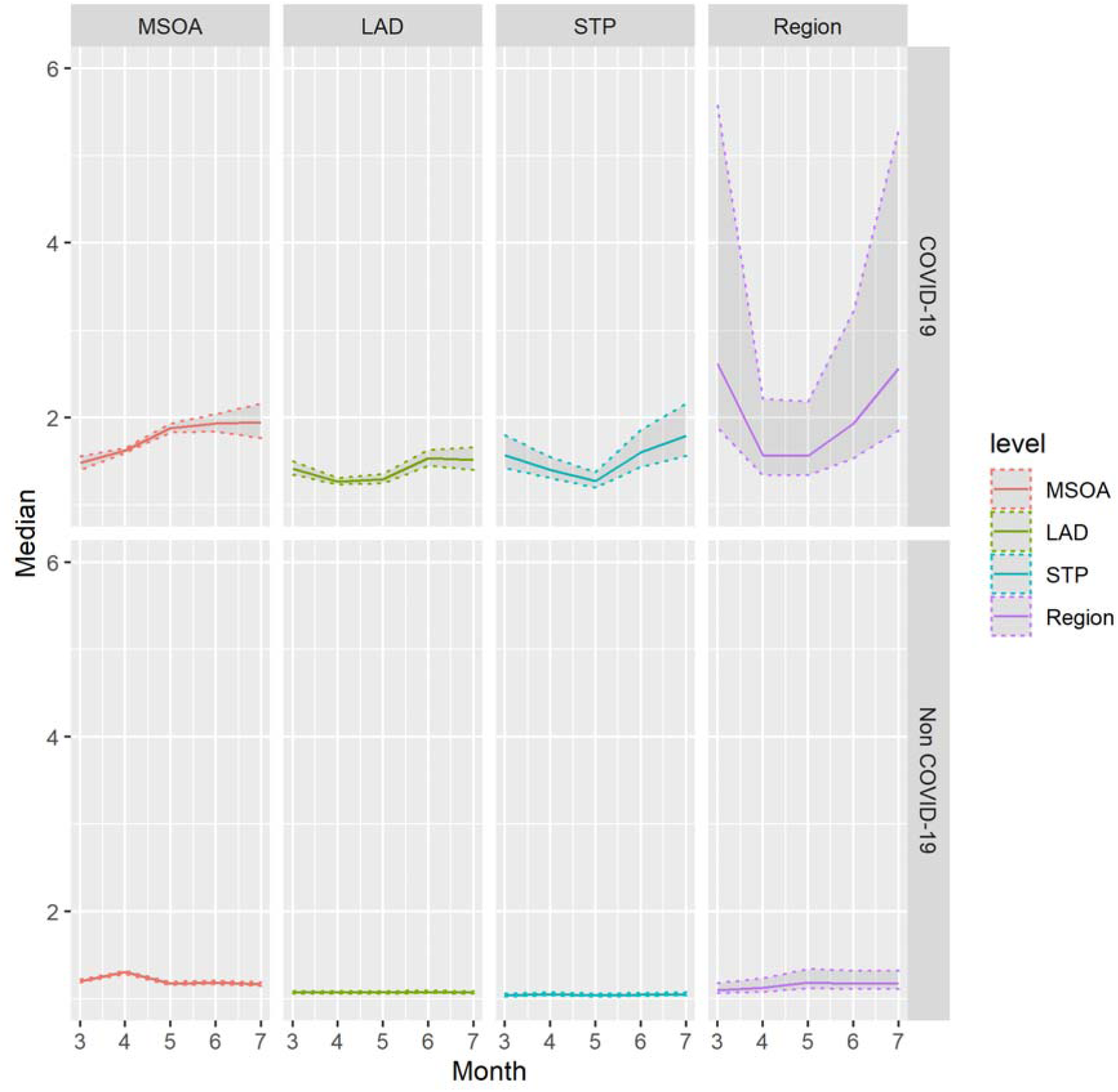
(top) and 2B (bottom) - Estimates of Median COVID-19 (top) and Non-COVID (bottom) Median Mortality Rate Ratio across spatial scales, by month from March to July 2020. Shaded areas indicate 2.5th and 97.5th percentile credible intervals of posterior parameter distributions.

Figure 2A illustrates the development over time of MRRs across different spatial scales. Greatest inequalities are expressed at the MSOA and Region levels. For instance, the COVID-19 Mortality MRR in July for MSOA is 1.96 (95% CI 1.76 – 2.18), suggesting within-LAD, between-MSOA differences typically imply a doubling of the rate of COVID-19 mortality. We see considerable temporal heterogeneity, inequality in COVID deaths in March is mostly at the regional level (MRR 2.59, 1.84-5.52), however this declines rapidly in April as mortality rates increase, with most inequality seen within-LAD between-MSOA. From April and May onwards, there is an increase in regional inequality as higher-level inequalities start to increase once again peaking again in July (MRR 2.53, 1.81-5.41).

The contrast with Figure 2B is stark. Mortality rates for non-COVID mortality are higher than those for COVID-mortality, they are far more spatially random, the highest rate ratio between areas at any of the scales is at MSOA level in April. Inequality over and above that predicted by stochastic variation is only seen in April for non-COVID mortality.

Spatial inequalities in COVID-19 mortality have clearly not been temporally stable, but we are also interested in whether the residual patterning of mortality is consistent over time. That is, whether regions with high mortality in March have high mortality in subsequent months. Figure 3 illustrates the correlation between area-level residuals from March-July.

**Figure 3:**
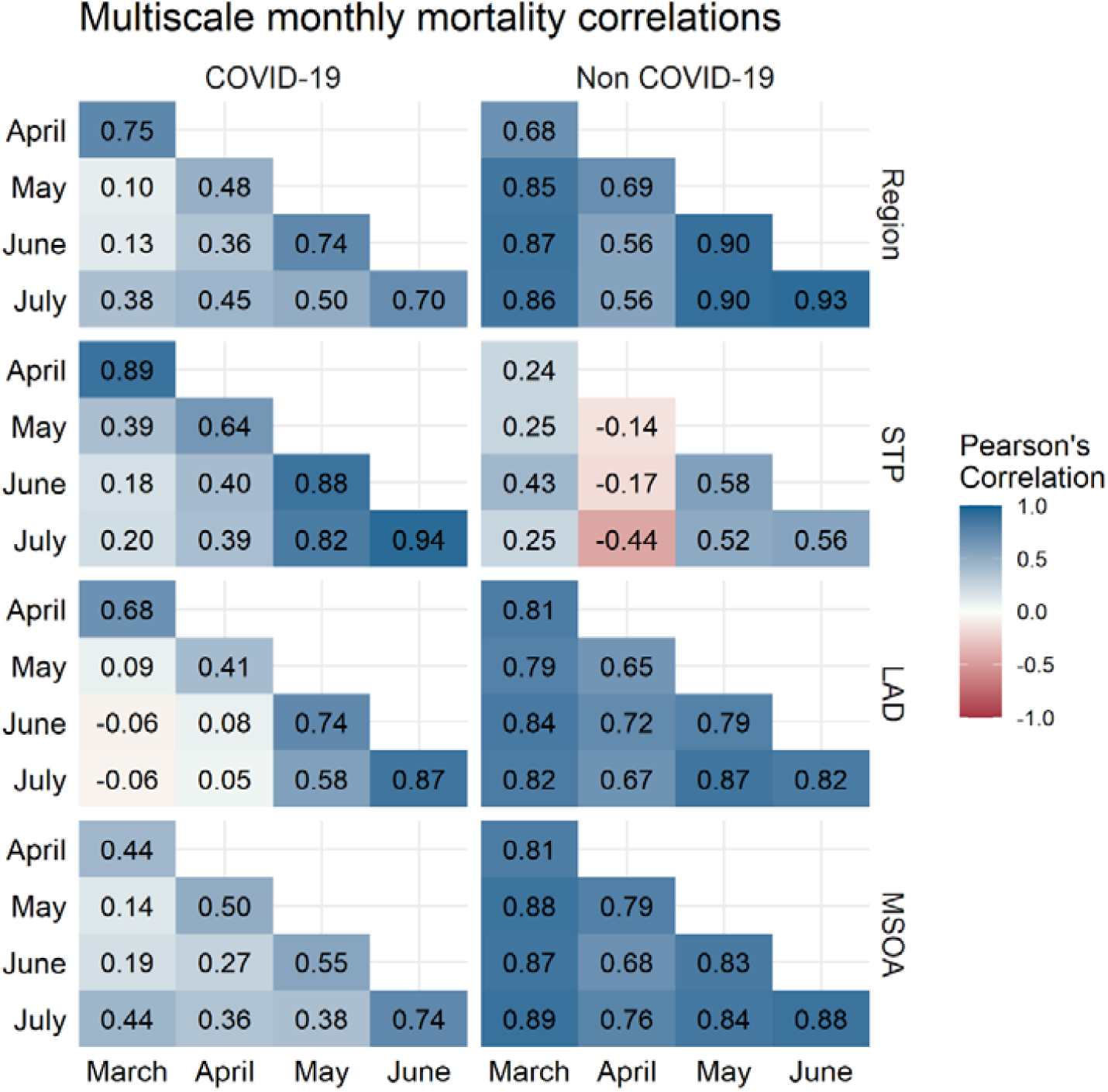
Correlation between month-specific structured residuals at 4 spatial scales for COVID-19 mortality (left) and non-COVID mortality (right). Colour indicates magnitude of correlation.

Correlations are broadly larger between adjacent months. However, this is not temporally consistent and area-level trends start to emerge, for instance, the highest correlation is between June and July for all levels for COVID-mortality. Excluding STP, correlations for non-COVID deaths are uniformly high, suggesting that the small amount of clustering we see in non-COVID mortality is highly temporally autocorrelated. STP-level non-COVID mortality residuals are low, and suggest unusual spatial patterning in April. MSOA COVID-mortality correlations increase consistently over the study period.

Having established spatial patterns of mortality inequality, we investigate to what degree these inequalities are explained by contextual deprivation. Being in the North, number of care homes and decomposed IMD estimates were all interacted with month. Results from fully adjusted models are displayed in Figures 4 and 5. Fixed effects estimates can be seen in Supplementary Table 2.

**Figure 4:**
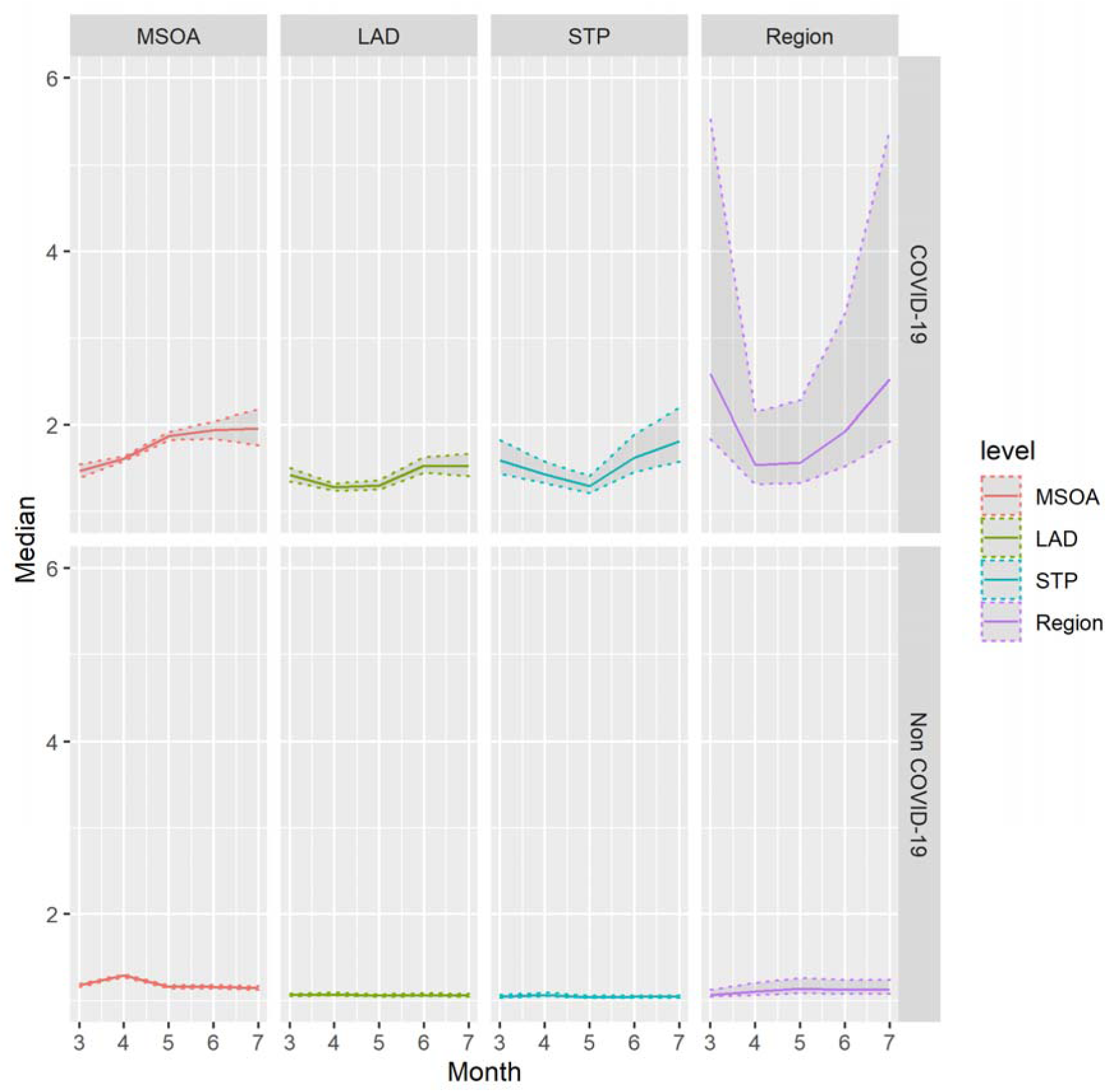
(top) and 4B (bottom) - Estimates of Median COVID-19 (4A) and Non-COVID (4B) Median Mortality Rate Ratio across spatial scales after inclusion of local area deprivation, by month from March to July 2020. Shaded areas indicate 2.5th and 97.5th percentile credible intervals of posterior parameter.

**Figure 5:**
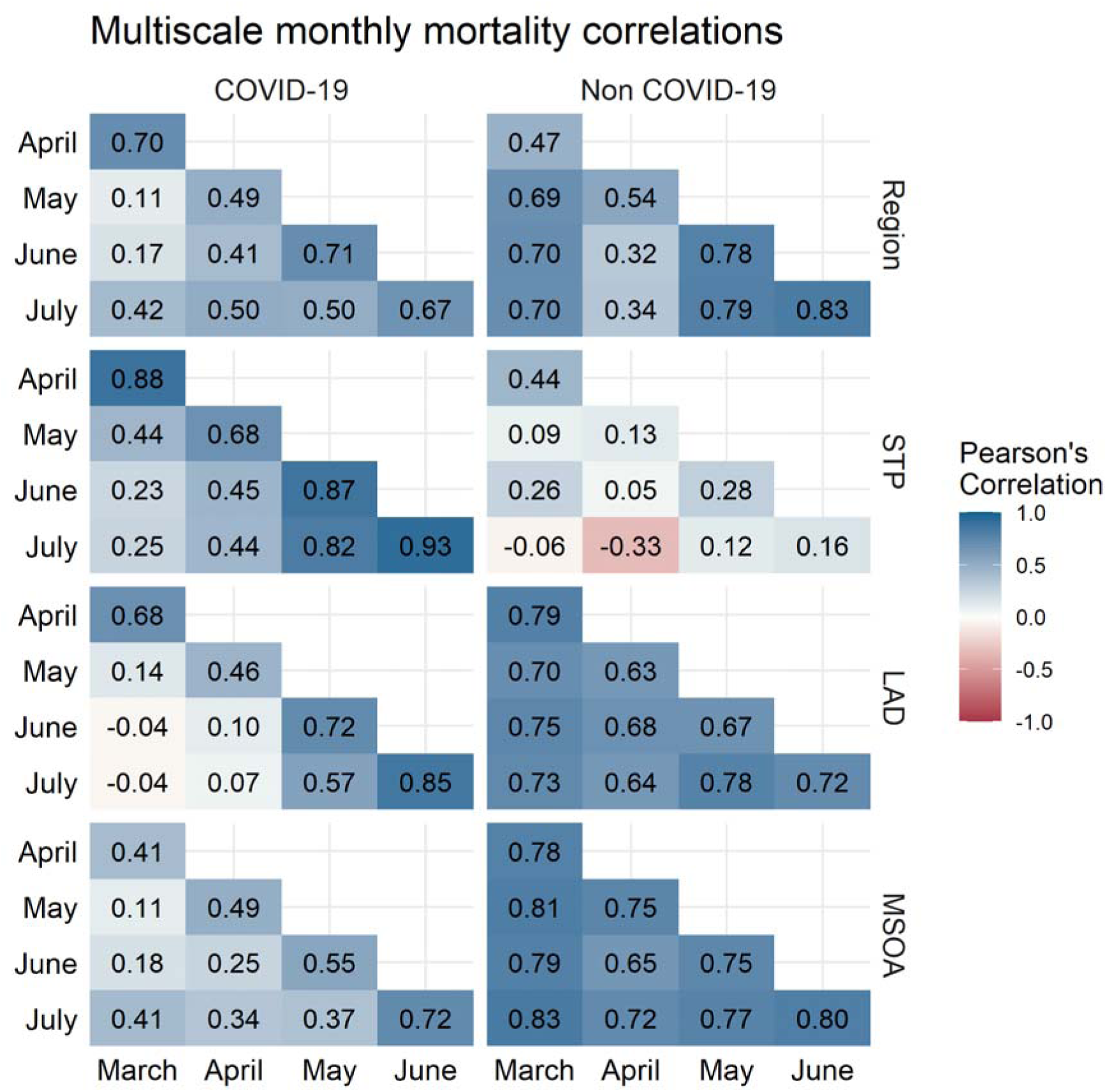
Correlation between month-specific structured residuals at 4 spatial scales for COVID-19 mortality (left) and non-COVID mortality (right) after adjusting for local deprivation. Colour indicates magnitude of correlation.

Figure 4 demonstrates that introducing IMD does not substantively change the variance structure of the model. In July MRRs are still large, with a regional MRR of 2.52 (1.81-5.40) and an MSOA MRR of 1.96 (1.76-2.18). The correlation plots presented in Figure 5 are similarly consistent with plots prior to the introduction of area-deprivation.

To test whether higher-level regional mortality residuals are a function of London’s abnormality, fully-adjusted models were rerun with London omitted, these are presented in Supplementary Analysis 3. Results suggest that some but not all higher-level regional inequality is driven by the abnormally high age-standardised mortality in London. Omitting London reduces the regional inequality to a similar order of magnitude to that of STP and LAD. Omitting London also suggests that some of the temporal autocorrelation in regional non-COVID-19 mortality is driven by London, with smaller correlations in Model SA1 than 2B.

## Conclusions

Our results highlight that spatial inequality in COVID-19 deaths at all scales is substantially higher than deaths due to other causes. Clearly, geography matters for an individual’s chance of dying of COVID-19 and it matters more than it does for dying of other causes.

The extent to which geography matters has changed over time. After April, the chance of someone dying of COVID-19 varied by where they lived to an increasing extent until July. We propose these changes likely reflect structural factors affecting risk of exposure or infection for two reasons. Firstly, phylogenetic evidence suggests aetiological evolution over the summer of 2020 impacted viral transmission, not disease progression (du Plessis et al. 2021; Volz et al. 2021). Thus, conditional on age structure, we anticipate mortality changes likely relate to changes in infection, not prognosis. Secondly, suppression measures make it unlikely that compositional migration patterns drove short term mortality differences.

The geography of COVID-19 mortality is structured at different scales. While this is often implicitly acknowledged in debate, particularly around north-south divides, we believe this is the first study to attempt to quantify and apportion this. We find inequalities at each examined scale but find particularly large inequalities at the largest (GOR) and smallest (MSOA) scales. Our results are consistent with other findings on clustered introduction of the virus producing large initial spatial inequalities, driven partially by high mortality in London. After March regional inequalities decline as COVID-19 mortality increases, before increasing in June and July with declining mortality rates. Excluding London, our findings remain consistent, with median comparisons between regions giving a two-fold mortality risk by July (MRR=2.16, 1.59 – 4.41).

Consistent with previous studies, deprived areas have greater COVID-19 mortality than non-deprived areas. There is some evidence supporting initial misclassification of COVID-19 mortality, as MSOA non-COVID-19 MRR pre-empts that of the COVID-19 MRR before declining, and care-homes predicting non-COVID mortality in April, prior to the care-home spike in COVID-19 mortality in May. There is strong evidence of emerging regional trends and a north-south divide by July. Moreover, such inequalities are not explained by deprivation, care homes or age structure. Results suggest that a devolved geographical approach to COVID-19 support is likely useful. This approach must be multiscale with local community strategies embedded within regional frameworks and must consider pre-existing health and social inequalities.

Our study has several limitations. Our model considers geography in a strictly hierarchical sense, where we know in which LAD/Region each MSOA/LAD is situated. It does not consider spatial contiguity/proximity and spatial networks. Clearly for infectious diseases, connections between and within areas are important and may explain some of the patterns we see here^2^.

We propose that our study informs outcomes beyond COVID-19, as the modelling approach is of clear utility for studying other health outcomes. Firstly, our approach can help advise where and how to apportion funding for tackling nationwide health inequalities. Assuming our goal is to reduce absolute inequalities, this approach informs how we might effectively do so by providing a structured framework for prioritising health inequalities between regions; between cities, towns or areas within regions; or between neighbourhoods within these cities, towns and areas.

Secondly, the modelling approach aims explicitly to distinguish policy-relevant systemic differences from chance variation. Models necessarily require assumptions about the degree of stochasticity which truly represents uncertainty, indeed phylogenetic evidence suggests UK SARS-CoV-2 evolution is a truly stochastic process (du Plessis et al. 2021). Spatial epidemiology, however, often fails to explicitly recognise chance variation as inherent in population healthx (Davey Smith 2011).

Latent structural information contained in spatial data is being deployed across a wide range of geographical contexts to investigate spatial inequalities (e.g. Bilal et al. 2020; Gibertoni et al. 2021; Otitoloju et al. 2020; Sannigrahi et al. 2020; Snyder and Parks 2020). Moreover these data are providing important evidence in establishing effects of associated structural exposures (Carmo et al. 2020; Daras et al. 2020; Fortaleza et al. 2020). Our method is readily extendable to such contexts in the presence of; existing, administrative; or bespoke, researcher-imposed higher-level spatial identifiers.

Continued monitoring of area-level mortality will be a fundamental piece of the evidence base helping mitigate the impacts of the COVID-19 pandemic. Mortality data at MSOA level is currently only available for the defined study period but analysing multi-scale inequalities in mortality is critical to evaluating the impact and efficacy of viral suppression measures. We suggest multiscale spatial inequalities which inform COVID-19 mortality, predominantly via exposure and infection, likely represent a key potential structural intervention for virus suppression.

## Data Availability

ONS COVID-mortality data is available here: https://www.ons.gov.uk/peoplepopulationandcommunity/birthsdeathsandmarriages/deaths/datasets/deathsinvolvingcovid19bylocalareaanddeprivation

ONS Administrative geographical identifiers are available here: https://geoportal.statistics.gov.uk

ONS population health estimates are available here: https://www.ons.gov.uk/peoplepopulationandcommunity/populationandmigration/populationestimates/datasets/middlesuperoutputareamidyearpopulationestimates.

ESRI care home estimates are available here: https://covid19.esriuk.com/datasets/e4ffa672880a4facaab717dea3cdc404_0

Equivalised UK IMD estimates are available here: https://data.bris.ac.uk/data/dataset/1ef3q32gybk001v77c1ifmty7x

2019 England IMD estimates are available here: https://www.gov.uk/government/statistics/english-indices-of-deprivation-2019

All code to reproduce analyses is available here: www.github.com/zimbabwelsh/covid_mort_spatseg

## Data Availability

ONS COVID-mortality data is available here: *https://www.ons.gov.uk/peoplepopulationandcommunity/birthsdeathsandmarriages/deaths/datasets/deathsinvolvingcovid19bylocalareaanddeprivation*

ONS Administrative geographical identifiers are available here: *https://geoportal.statistics.gov.uk*

ONS population health estimates are available here: *https://www.ons.gov.uk/peoplepopulationandcommunity/populationandmigration/populationestimates/datasets/middlesuperoutputareamidyearpopulationestimates.*

ESRI care home estimates are available here: *https://covid19.esriuk.com/datasets/e4ffa672880a4facaab717dea3cdc404_0*

Equivalised UK IMD estimates are available here: *https://data.bris.ac.uk/data/dataset/1ef3q32gybk001v77c1ifmty7x*

2019 England IMD estimates are available here: *https://www.gov.uk/government/statistics/english-indices-of-deprivation-2019*

All code to reproduce analyses is available here: *www.github.com/zimbabwelsh/covid_mort_spatseg*

## Acknowledgements

GG is supported by an ESRC postdoctoral fellowship [ES/T009101/1].

## Author Contributions

G.G. and G.O. conceived the research

G.G. performed the analysis

G.G. and G.O. generated figures for final manuscript

All authors discussed results, sensitivity analyses and contributed to the final manuscript.

## Ethics Statement

This research was carried out entirely using aggregate unidentifiable data made available by the UK Office for National Statistics, Environmental Systems Research Institute, or in publicly available datasets from previous research. No individual-level data was used, and no participants were involved in this research.

## Footnotes

1 Area level resource prioritisation is common practice in the UK. https://assets.publishing.service.gov.uk/government/uploads/system/uploads/attachment_data/file/466509/Resource_allocation_for_public_health.pdf

2 Sensitivity analysis replicated our final models using TTWAs and did not alter the higher-level structure of model results (see Supplementary Analysis 1).

