## Supplementary Materials for "Interrogating structural inequalities in COVID-19 Mortality in England and Wales"

*Model Specification*

Equation 1, below, sets out our approach. For clarity this is illustrated with a simpler version of the model with only two months (March and April) and two geographic scales (MSOA and LAD). In practice we include 5 months and 4 geographical scales as the equation below is readily extendable . Although the data for this model have two levels, MSOA and LAD, the specified model has an extra pseudo-individual level, level 1, though in reality the units at this level are identical to the MSOAs at level 2. The reason for this extra level is it enables us to include two sources of variation in the model; stochastic variation in the counts following a Poisson distribution and the overdispersion identified above on top of this which we consider to be systematic inequality (Leckie et al. 2020). The variance at level 1 is constrained to be equal to the underlying rate for each group, as in a Poisson distribution, while the extra-Poisson variance at level 2 is then estimated in the model giving a measure of systematic inequality at the MSOA level.

$$O_{ijk} \sim Poisson\left( \pi_{ijk} \right)$$

$${Log}_{e}\pi_{ijk}={Log}_{e}\left( E_{ijk} \right)+ \beta_{1jk}March+ \beta_{2jk}April$$

$$\beta_{1jk}= \beta_{1}+ v_{1k}+u_{1jk}$$

$$\beta_{2jk}= \beta_{2}+ v_{2k}+u_{2jk}$$

$$\left[ \begin{matrix} v_{1k} \\ v_{2k} \end{matrix} \right] \sim N \left( 0,\left[ \begin{matrix} \begin{matrix} {\sigma^{2}}_{v1} \end{matrix} & \\ \sigma_{v12} & {\sigma^{2}}_{v2} \end{matrix} \right] \right)$$

$$\left[ \begin{matrix} u_{1jk} \\ u_{2jk} \end{matrix} \right] \sim N \left( 0,\left[ \begin{matrix} \begin{matrix} {\sigma^{2}}_{u1} \end{matrix} & \\ \sigma_{u12} & {\sigma^{2}}_{u2} \end{matrix} \right] \right)$$

$$Var(O_{ijk}|\pi_{ijk})$$

In the model O_ijk is a long stacked vector of observed counts for individual, i in MSOA, j, in Local Authority District, k with repeated observations for each month in the model. The observed counts are treated as coming from a Poisson distribution with mean rate, π_ijk. The natural log of this underlying rate is modelled.

March and April are dummy variables which indicate which month each count belongs to. E_ijk is the expected count for each MSOA in each month if deaths per unit of population were evenly distributed across MSOAs, or in other words if there was no inequality. The natural log of this is included in the model as an offset, with a coefficient constrained to 1. β1 and β2 give the log average (median) rate across MSOAs for March and April respectively.

Returning to the description of the model $u_{1jk}$ and $u_{2jk}$are, therefore, differences from the expected rate for MSOA$j$ for March and April respectively, while $v_{1k}$ and $v_{2k}$are the same the same for LADslevel. Positive values denote a higher than expected log rate for that geographical unit, while negative values denote a lower than expected log rate. These differences at each level are assumed to come from a joint normal distribution and can be summarized by variances $\begin{matrix} {\sigma^{2}}_{u1} \end{matrix}$, $\begin{matrix} {\sigma^{2}}_{u2} \end{matrix}$ , $\begin{matrix} {\sigma^{2}}_{v1} \end{matrix}$, and $\begin{matrix} {\sigma^{2}}_{v2} \end{matrix} ,$respectively. It is these variances which measure the inequality and are the parameters of interest. For example ${\sigma^{2}}_{u1}$is a measure of inequality for March at the MSOA level.

Finally, $\sigma_{u12}$ and $\sigma_{v12}$are the covariances, between the two groups at the two levels which can be standardised to give a correlation ranging between -1 and 1. These allow us to see if it is the same MSOAs and LADs which have high deaths in March are also the same MSOAs which have high deaths in April.

*Supplementary Analysis*

| *ONS Reported Mortality* | COVID-19 Mortality | Non-COVID-19 Mortality |
| --- | --- | --- |
| March | 4759 | 48 563 |
| April | 30 634 | 50 611 |
| May | 11 932 | 38 114 |
| June | 3391 | 34 772 |
| July | 1052 | 35 097 |

Supplementary Table 1: Monthly Mortality Counts for COVID-19 and Non-COVID-19 causes of death.

Fixed effect results allow us to investigate more clearly the following two questions of interest:

1. Is the effect of local-area deprivation consistent both within and between higher level spatial units?
2. Does an observed north-south mortality divide exist net of deprivation and net of age-structure/care homes?

Supplementary Table 1: Fixed Effect Coefficients for Models 2A and 2B, reflecting mortality risk ratios for COVID-19 and Non COVID-19 mortality.

|  | 2A: COVID-19 Mortality | | | 2B: Non-COVID-19 Mortality | | |
| --- | --- | --- | --- | --- | --- | --- |
|  | Rate Ratio | 2.5% CI | 97.5% CI | Rate Ratio | 2.5% CI | 97.5% CI |
| MSOA % 25-44 | 1.001 | 0.996 | 1.006 | 1.002 | 1.000 | 1.004 |
| MSOA % 45-64 | 1.022 | 1.015 | 1.028 | 1.022 | 1.020 | 1.025 |
| MSOA % 65-74 | 0.945 | 0.933 | 0.957 | 0.985 | 0.980 | 0.990 |
| MSOA % 75+ | 1.123 | 1.112 | 1.135 | 1.091 | 1.087 | 1.096 |
| North*Mar | 0.366 | 0.041 | 3.604 | 1.119 | 0.959 | 1.308 |
| North*Apr | 0.700 | 0.295 | 1.859 | 1.086 | 0.851 | 1.405 |
| North*May | 1.623 | 0.649 | 5.038 | 1.260 | 0.960 | 1.677 |
| North*Jun | 3.077 | 0.802 | 16.184 | 1.238 | 0.947 | 1.581 |
| North*Jul | 3.277 | 0.462 | 30.753 | 1.258 | 0.965 | 1.594 |
| Carehomes*Mar | 1.020 | 1.006 | 1.034 | 1.039 | 1.035 | 1.042 |
| Carehomes*Apr | 1.062 | 1.053 | 1.069 | 1.058 | 1.054 | 1.063 |
| Carehomes*May | 1.082 | 1.071 | 1.093 | 1.037 | 1.033 | 1.042 |
| Carehomes*Jun | 1.051 | 1.035 | 1.067 | 1.032 | 1.028 | 1.037 |
| Carehomes*Jul | 1.030 | 1.005 | 1.057 | 1.028 | 1.023 | 1.032 |
| RegIMD*Mar | 1.592 | 0.362 | 4.938 | 1.009 | 0.932 | 1.095 |
| RegIMD*Apr | 1.401 | 0.839 | 2.217 | 1.029 | 0.898 | 1.169 |
| RegIMD*May | 1.057 | 0.551 | 2.044 | 0.961 | 0.828 | 1.117 |
| RegIMD*Jun | 0.839 | 0.351 | 1.813 | 0.976 | 0.856 | 1.131 |
| RegIMD*Jul | 0.757 | 0.205 | 2.113 | 0.963 | 0.846 | 1.111 |
| L-RIMD*Mar | 1.099 | 1.033 | 1.169 | 1.063 | 1.049 | 1.078 |
| L-RIMD*Apr | 1.068 | 1.027 | 1.110 | 1.045 | 1.030 | 1.062 |
| L-RIMD*May | 1.010 | 0.967 | 1.055 | 1.055 | 1.041 | 1.070 |
| L-RIMD*Jun | 1.005 | 0.936 | 1.080 | 1.055 | 1.040 | 1.071 |
| L-RIMD*Jul | 1.037 | 0.946 | 1.135 | 1.050 | 1.035 | 1.064 |
| M-LIMD*Mar | 1.097 | 1.062 | 1.134 | 1.116 | 1.105 | 1.128 |
| M-LIMD*Apr | 1.121 | 1.100 | 1.142 | 1.107 | 1.094 | 1.120 |
| M-LIMD*May | 1.100 | 1.071 | 1.129 | 1.107 | 1.094 | 1.119 |
| M-LIMD*Jun | 1.096 | 1.053 | 1.141 | 1.112 | 1.099 | 1.125 |
| M-LIMD*Jul | 1.114 | 1.045 | 1.188 | 1.104 | 1.091 | 1.117 |

Supplementary Table 2 presents the fixed effect estimates from models 2A and 2B. We are primarily interested in the estimates for North and for IMD. Estimates are imprecise, but there is some evidence that the effect of living in the North has flipped from initially slightly protective to risk-inducing over the study period. Deprivation does not appear to have a consistent association with mortality over different spatial scales. The effect of regional IMD is imprecisely estimated, but seems to flip over the course of the study period, with this effect estimated net of being in the North. There is strong evidence that high MSOA deprivation consistently predicts excess death across both COVID and non-COVID mortality. However, between LAD deprivation matters consistently for non-COVID, but diminishes greatly for COVID-19 mortality from May-July. Care homes consistently predict excess COVID-19 mortality, peaking in May. Care homes most strongly predict non-COVID death in April before declining. This, taken with the negative residual correlation between April Non-COVID deaths and other months may indicate misclassification in COVID-19 deaths in care homes during April.

*Supplementary Analysis 1*

*Model SA1*, Replicating Model 2A with TTWAs replacing LADs

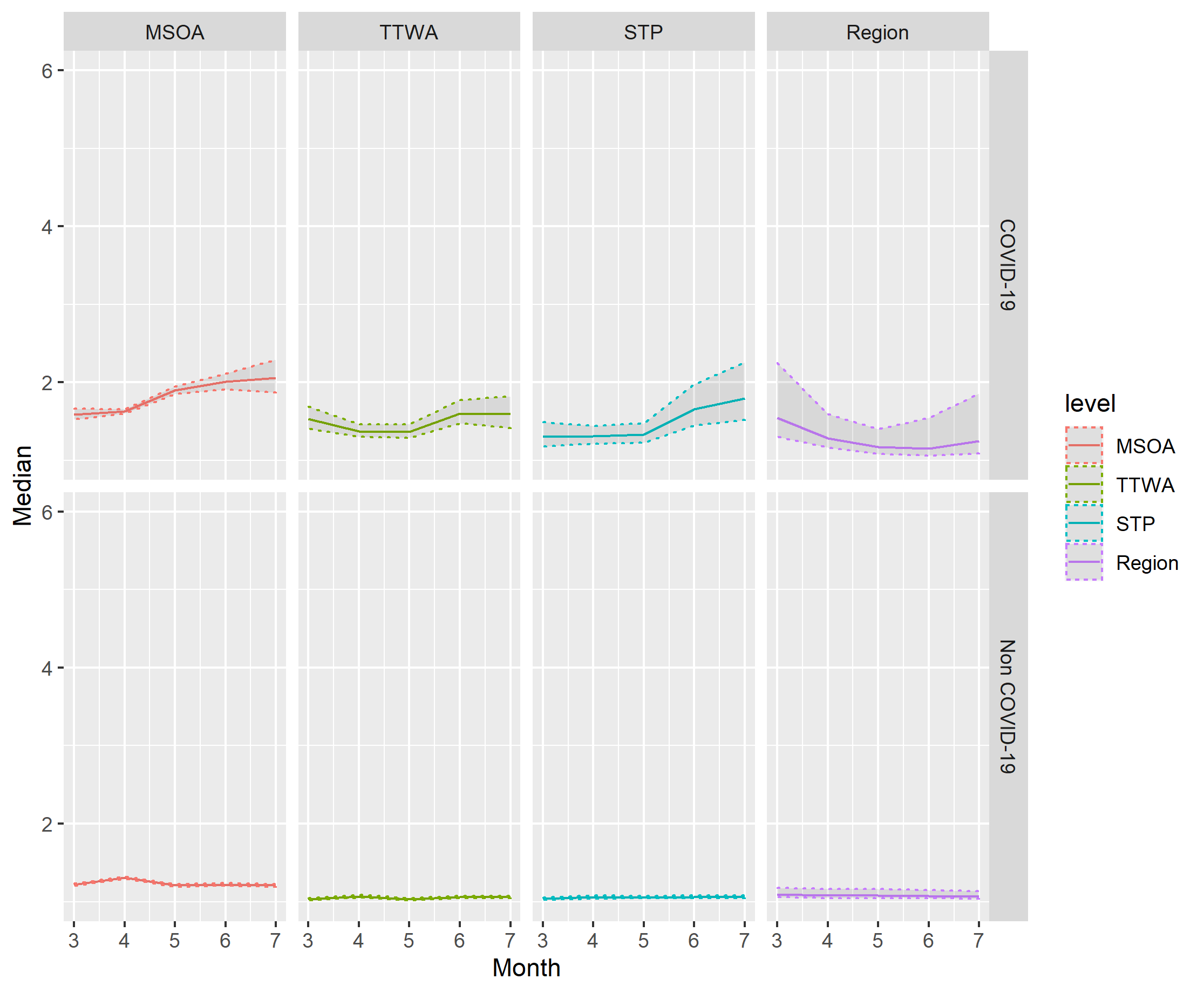

Supplementary Figure 1A (top) and 1B (bottom): Estimates of Median COVID-19 (1A) and Non-COVID (1B) Median Mortality Rate ratio including TTWA instead of LAD across spatial scales after inclusion of local area deprivation. Shaded areas indicate 2.5th and 97.5th percentile credible intervals of posterior parameter.

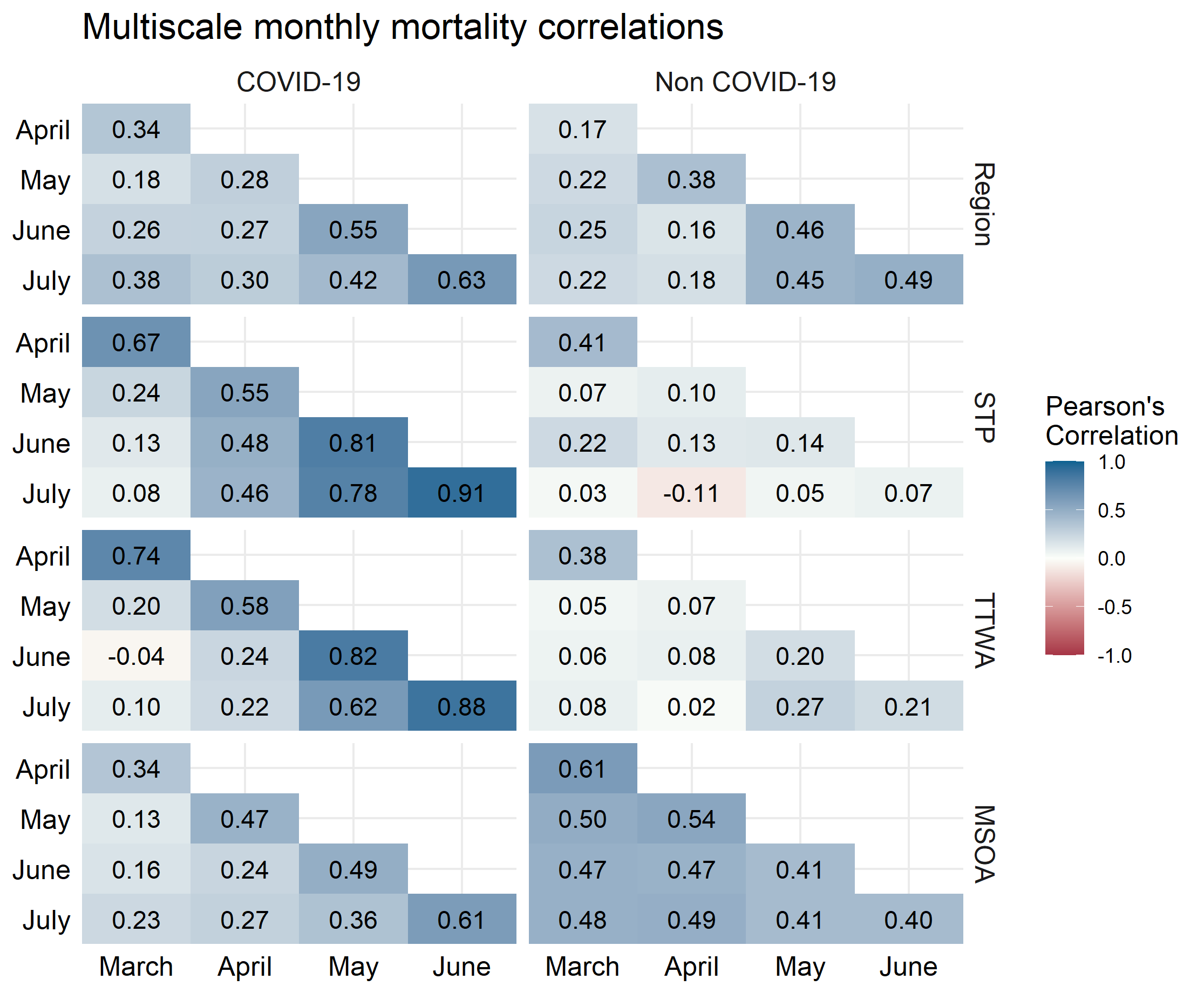

Supplementary Figure 2: Correlations between month-specific structured residuals at 4 spatial scales for COVID-19 mortality (left) and non-COVID mortality (right) after adjusting for local deprivation, omitting region of London.

*Supplementary Analysis 2*

*Model SA2*, Replicating Model 2, excluding Wales and using 2019 England IMD instead of 2015 UK-wide IMD

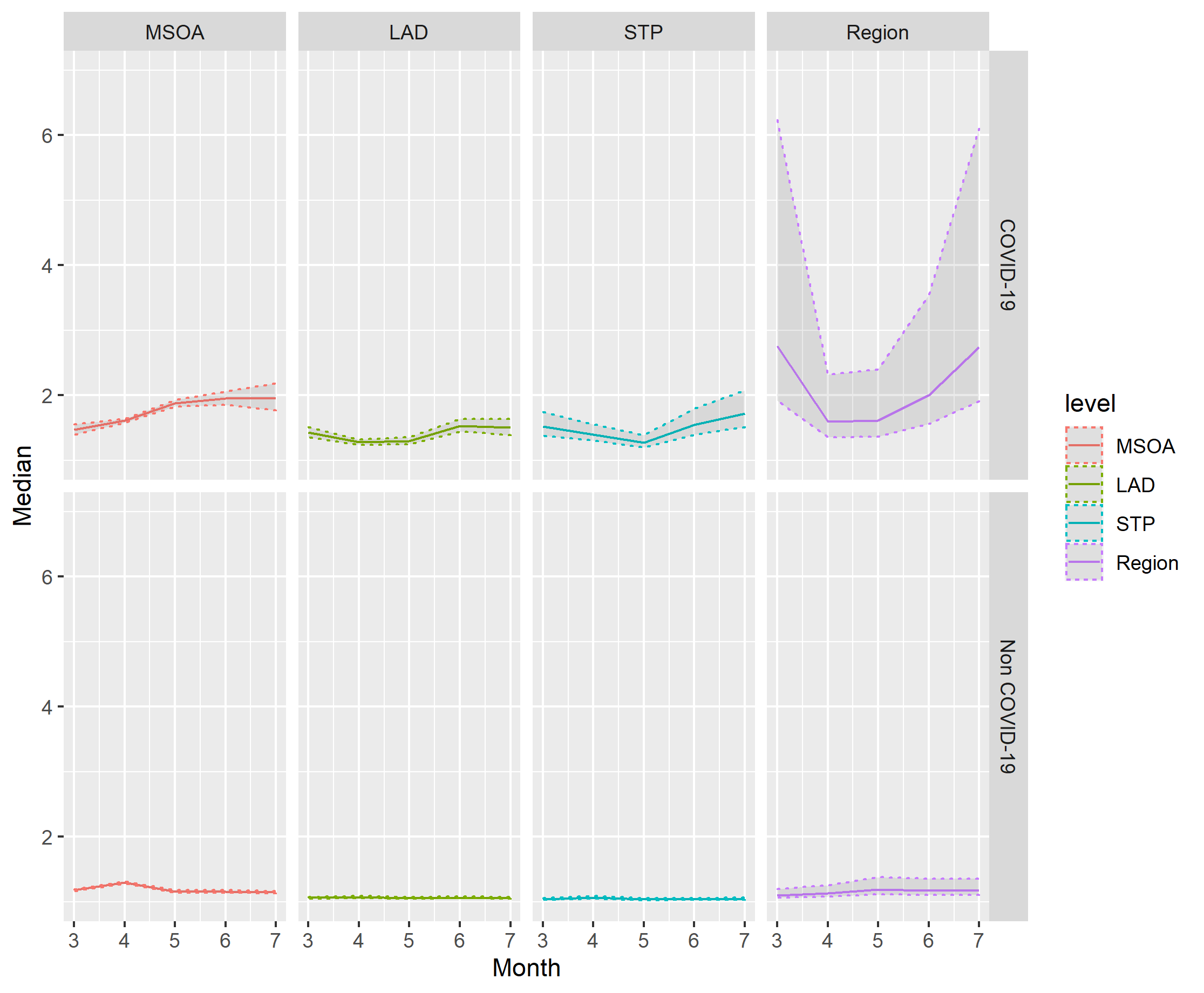

Supplementary Figure 3A (top) and 3B (bottom): Estimates of Median COVID-19 (3A) and Non-COVID (3B) Median Mortality Rate Ratio across spatial scales after exclusion of Wales and inclusion of 2019 IMD. Shaded areas indicate 2.5th and 97.5th percentile credible intervals of posterior parameter.

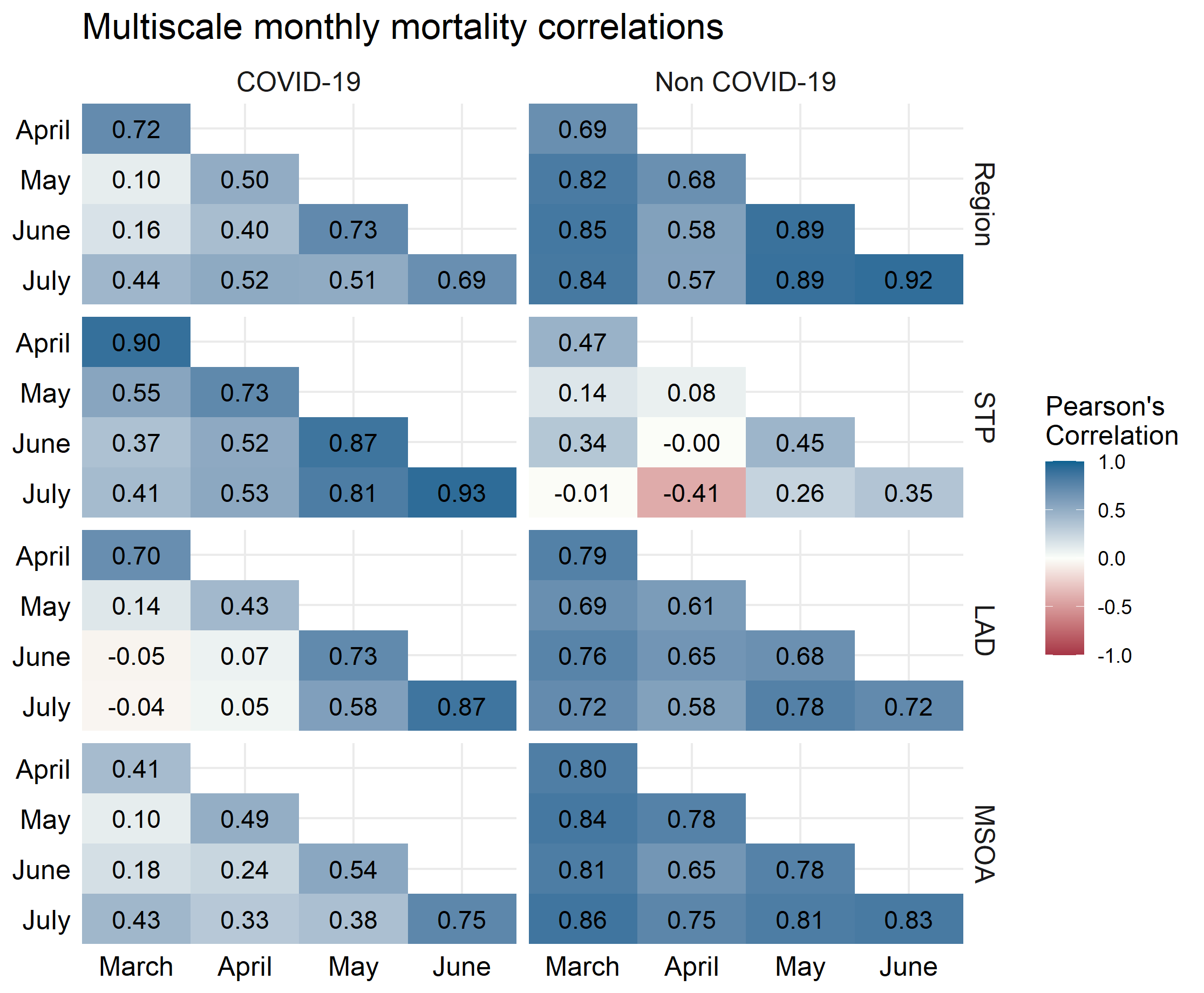

Supplementary Figure 4: Correlations between month-specific structured residuals at 4 spatial scales for COVID-19 mortality (left) and non-COVID mortality (right) after adjusting for 2019 IMD, omitting Wales.

*Supplementary Analysis 3*

*Model SA3* – Replicating Model 2, with London omitted from results:

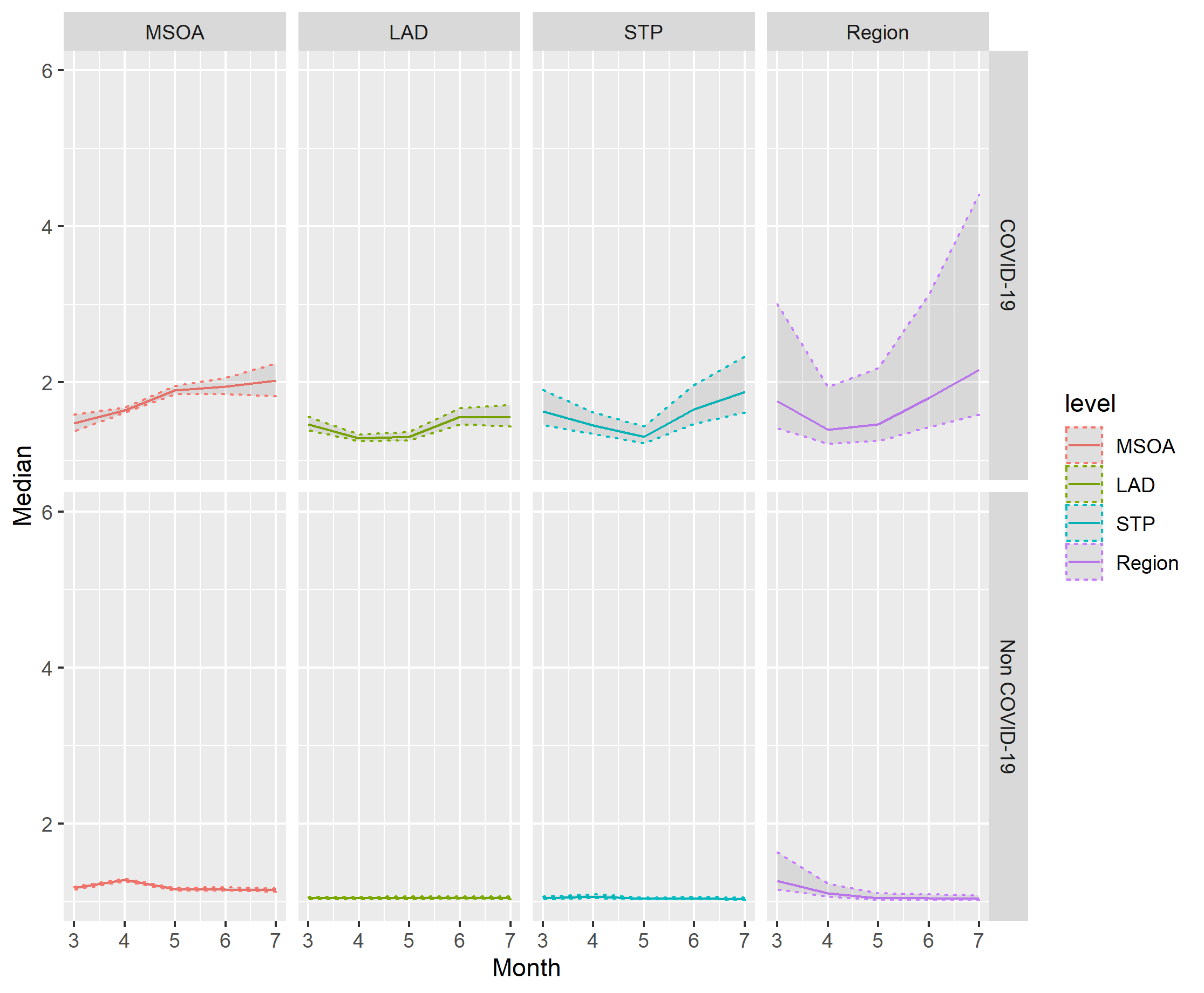

Supplementary Figure 5A (top) and 5B (bottom): Estimates of Median COVID-19 (5A) and Non-COVID (5B) Median Mortality Rate Ratio omitting region of London across spatial scales after inclusion of local area deprivation. Shaded areas indicate 2.5th and 97.5th percentile credible intervals of posterior parameter.

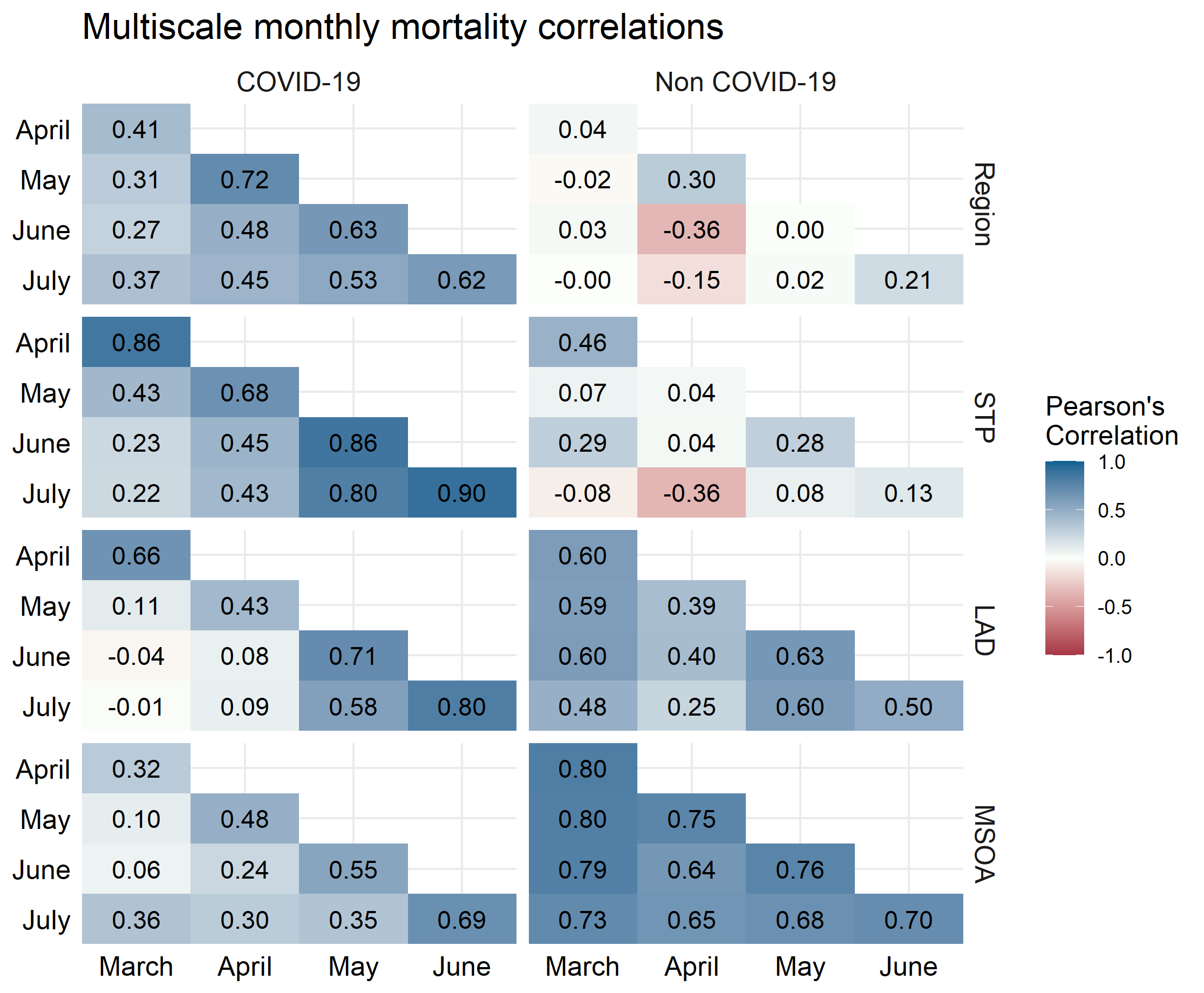

Supplementary Figure 6: Correlations between month-specific structured residuals at 4 spatial scales for COVID-19 mortality (left) and non-COVID mortality (right) after adjusting for local deprivation, omitting region of London.
